# Predicting HPV/p16 Status and Molecular Subtypes from Histopathology Slides using Deep Learning in Vulvar Squamous Cell Carcinoma

**DOI:** 10.64898/2026.10.22.26366203

**Authors:** Nikki J. van den Berg, Willemijn L. van der Kolk, Kim Landsbergen, Kim Kortekaas, Linda Nooij, Anne Fleur van Velzen, Nienke C. te Grootenhuis, the AIRMEC Consortium, Nanda Horeweg, Lena C. van Doorn, Maaike H.M. Oonk, Jos Bart, Tjalling Bosse

## Abstract

Vulvar squamous cell carcinoma (VSCC) comprises three prognostically distinct molecular subtypes: human papillomavirus (HPV)-associated, HPV-independent/p53-wildtype, and HPV-independent/p53-abnormal. Classification requires p16/p53 immunohistochemistry, which consumes tissue and is often incomplete in retrospective cohorts. We investigated whether deep learning could predict HPV/p16 status and molecular subtypes directly from routine hematoxylin and eosin–stained whole-slide images. Using features extracted with H-optimus-1, we developed two attention-based multiple-instance learning models: HERA, predicting HPV/p16 status, and HERMES, predicting the three molecular subtypes. Models were developed on 644 patients from three Dutch VSCC cohorts and tested on 286 patients from two independent cohorts. In independent testing, HERA achieved an area under the curve (AUC) of 0.84 (95% CI 0.75–0.91), with specificity 0.93, negative predictive value 0.94, and sensitivity 0.68. HERA-predicted groups reproduced the prognostic separation of HPV testing (log-rank p<0.001). HERMES achieved a macro-average AUC of 0.70 (0.66–0.75), with good discrimination for HPV-associated (0.80) but limited separation of HPV-independent/p53-abnormal (AUC 0.68) and HPV-independent/p53-wildtype (AUC 0.63). HERMES-predicted classes reproduced the prognostic hierarchy, with 5-year recurrence rates of 14.0% (HPV-associated), 31.1% (HPV-independent/p53-wildtype) and 43.3% (HPV-independent/p53-abnormal) (p=0.0051). Highly attended tiles showed basaloid cells and koilocytotic-like atypia in HPV-associated tumors, keratinizing nests with desmoplastic stroma in HPV-independent/p53-abnormal tumors, and well-differentiated keratinocytes with spongiosis in HPV-independent/p53-wildtype tumors. HERA could assign HPV/p16 status to large retrospective VSCC cohorts lacking ancillary testing, without consuming tissue, enabling HPV-stratified outcome analyses. Before such use, external validation of its discrimination and calibration across scanners and populations is required. HERMES suggests that molecular subtype information is encoded in hematoxylin and eosin–stained slides, but requires improved distinction between the HPV-independent subtypes.

## Introduction

Vulvar squamous cell carcinoma (VSCC) is a rare malignancy with an incidence of 1–4 per 100,000 women each year.^1^ Its incidence is rising, partly due to increasing human papillomavirus (HPV) prevalence and population aging.^1^ Current diagnostic and treatment regimens are based on clinicopathologic factors, including tumor size, depth of invasion, and inguinofemoral lymph node status, as defined by the International Federation of Gynecology and Obstetrics (FIGO) staging system.^2^ VSCC arises through two distinct carcinogenic pathways, HPV-associated (HPV-a) and HPV-independent (HPV-i), which differ in their biological and clinicopathologic characteristics. The more recent subdivision of HPV-i tumors by p53 status yields three prognostic molecular subtypes: HPV-a, HPV-i with wildtype p53 (HPV-i/p53wt), and HPV-i with abnormal p53 (HPV-i/p53abn).^3^ HPV-a tumors generally show favorable prognosis, whereas HPV-i/p53abn tumors display aggressive behavior and have a higher risk of recurrence.^4^

Molecular classification is increasingly integrated into routine pathological reporting and is anticipated to assume an expanding role in prognostic assessment, treatment allocation, and the design of future clinical trials. Diagnostic tests for molecular stratification require immunohistochemical (IHC) and molecular assays; p16 overexpression is used as a surrogate marker for HPV infection, and p53 IHC serves as a surrogate marker for *TP53* mutation status.^5–7^ These ancillary tests require cutting unstained slides from formalin-fixed paraffin-embedded (FFPE) blocks, which are not always available in (older) retrospective cohorts for which one wants to infer HPV/p16 status for research purposes.

Recent advances in computational pathology have demonstrated that deep learning (DL) models can predict clinically relevant molecular alterations directly from routine H&E slides, indicating that genomic and viral-associated changes may be reflected in tumor morphology. In head and neck squamous cell carcinoma, a tumor type that resembles VSCC, several groups have reported HPV status prediction using DL with areas under the curve (AUCs) ranging from 0.77–0.84 in external validation.^8–10^ Although it is uncertain whether similar morphologic signatures exist in VSCC, it is plausible to hypothesize that HPV-associated morphology in VSCC may also be predictable by DL models.

Therefore, we investigated whether HPV/p16 status and molecular subtypes can be inferred directly from routine H&E whole-slide images (WSIs) using DL. First, to assess HPV/p16 status, we trained **HERA** (**H**&**E**-based **R**ecognition of HPV-**A**ssociated VSCC), a DL model that predicts the probability of HPV/p16 association (the “HERA-score”). Second, to predict the three molecular subtypes, we developed **HERMES** (**H**&**E**-based **R**ecognition of **M**olecular **E**xpressions and **S**ubtypes), which is trained to predict molecular subtypes directly from H&E slides. Beyond performance evaluation, we analyzed the morphologic features driving HERA and HERMES predictions which define the prototypic morphologies of HPV-a, HPV-i/p53wt, and HPV-i/p53abn VSCC. We further evaluated clinical outcome data to evaluate the prognostic value of DL classifications.

## Materials and methods

### Cohorts

The Erasmus Medical Center (EMC) cohort retrospectively included 328 VSCC patients from January 2000 to December 2015 at EMC, The Netherlands. The Leiden University Medical Center (LUMC)-I cohort retrospectively included 176 VSCC patients from January 2000 to December 2015 at the LUMC, The Netherlands. For both cohorts, the inclusion criteria were: primary VSCC, surgical treatment with curative intent and availability of FFPE blocks from resection samples.^3^ The patients were treated according to the Dutch national guideline: wide local excision or vulvectomy and a uni- or bilateral sentinel lymph node procedure and/or inguinofemoral lymphadenectomy, and if necessary adjuvant (chemo)radiotherapy.

The LUMC-II cohort retrospectively included 173 VSCC patients from 2016 to 2022 at LUMC, The Netherlands, with the following inclusion criteria: primary VSCC, surgical treatment with curative intent, no previous (chemo)radiotherapy in the pelvic area and with age older than 18 years.^11^ The patients were treated standard-of-care with curative intent: a radical local excision or vulvectomy, and a uni- or bilateral sentinel node procedure and/or inguinofemoral lymphadenectomy and/or adjuvant (chemo)radiotherapy.

The University Medical Center Groningen (UMCG)-I cohort retrospectively included 246 VSCC patients from 2020 to 2023 at UMCG, The Netherlands, with the following inclusion criteria: primary VSCC with a tumor size < 4 cm, depth of invasion >1 mm, not encroaching surrounding structures, such as the anus, vagina and urethra. The patients were treated with a radical local excision of the tumor and underwent a sentinel node procedure. In case of sentinel node metastasis, patients underwent additional treatment with radiotherapy and/or inguinofemoral lymphadenectomy.

The UMCG-II cohort retrospectively included 153 vulvar carcinoma patients from 2000 to 2007 at the University Medical Center Groningen (UMCG), The Netherlands, with the following inclusion criteria: unifocal primary disease with a depth of invasion > 1 mm, primary treatment (without prior neoadjuvant chemo- and/or radiotherapy), and treatment with curative intent.^12,13^

### Datasets

Data from five cohorts were assembled, leading to a total of 1,076 VSCC patients at start (Figure S1). First, 66 patients were excluded from the LUMC-I and EMC cohorts due to either previous (chemo)radiotherapy in the pelvic area (n=25), no curative intent (n=1), or no FFPE block available (n=40). Then, HPV-i VSCC patients with missing p53 IHC or missing *TP53* status were excluded (n=12). Also, patients with no VSCC in the FFPE block at revision (n=1), patients with usual vulvar intraepithelial neoplasia at revision (n=2), no tumor in the available material (n=19), no material available for scanning (n=22), or no electronic patient record of the patient available (n=12) were excluded. In total, H&E-stained slides from 942 VSCC patients were included for scanning, after which patients were excluded for bad tissue quality (n=8) or failed scanning (n=4). This led to a total of 930 patients being included for analysis.

One representative WSI capturing the tumor as a whole was chosen for each patient. This was done by one experienced, expert gynecopathologist (TB) according to standard practice at pathology institutes worldwide. Model development used 644 VSCC WSIs in total (one slide per patient), including the LUMC-II (n=166), EMC (n=232), UMCG-I (n=246) cohorts. From this total, 20% was randomly sampled as an internal validation set, stratified by the three molecular subtypes. This internal validation set was used to compute the classification threshold. The remaining 80% was used for five-fold cross validation. Independent testing was done on 286 WSIs from the LUMC-I (n=147) and the UMCG-II (n=139) cohorts, due to available clinical outcome data and largely complete HPV-polymerase chain reaction (PCR) results.

H&E slides were digitized at 40× objective magnification on three scanner platforms: the P480 (3DHistech; LUMC-I, LUMC-II and EMC; 0.194 µm/px), the IntelliSite Ultra Fast Scanner (Philips; UMCG-I; 0.250 µm/px), and the Aperio ScanScope (Leica Biosystems; UMCG-II; 0.230 µm/px) (Table S1). Because native resolution differed between scanners, tiles were defined by physical size: non-overlapping tiles of 180 × 180 µm were extracted and resized to 224 × 224 pixels (≈0.80 µm/px) before feature extraction. Tiles with more than 20% white background were excluded. Tiles were ImageNet normalized.^14^

### Definitions of ground-truth labels

Ground-truth labels, the gold standard where DL predictions are compared to, were a composite of the results of p16 and p53 immunohistochemistry and HPV-PCR testing, as previously reported.^3,11–13^ Cases were labeled as HPV-a if HPV-PCR testing was positive with high-risk types 16, 18, 31, 33, 52, 53, 58, or when diffuse block-type p16 expression was found in case no HPV-PCR testing was performed.^15^ Cases were labeled as HPV-i when HPV-testing detected no, low risk or undetermined HPV-types (types 6, 11, 44, 54, and 70).^15^ Subsequently, to assign the molecular subtype to HPV-i cases: cases were labeled as p53abn if p53 IHC staining showed mutant patterns (Table S5).^6^ Cases were labeled as p53wt if p53 IHC showed wildtype patterns (Table S5).^6^ When HPV-PCR was unavailable, p16 IHC was used (n=383 (59.5%) in model development, n=1 (0.3%) in independent testing).^5^ In case p53 IHC pattern assessment was inconclusive and next generation sequencing data was available for *TP53* mutation status, the *TP53* mutation status was used (n=3 (1%) in independent testing).^7^ Therefore, the ground-truth labels represent a composite HPV/p16 status and *TP53*/p53 status rather than uniformly PCR-confirmed HPV and p53 IHC status.

### Development of HERA

HERA predicts HPV/p16 status (the “HERA-score”) from one WSI with representative VSCC tissue. To develop this model, we evaluated three foundation models (Virchow2, UNI2-h, H-optimus-1), network architectures (size of fully connected layers and the attention module) and training hyperparameters, that were optimized via five-fold cross-validation to maximize mean area under the receiver operating characteristic curve (AUC) (Supplementary Table S2).^16–18^ The optimal configuration used H-optimus-1 as foundation model, an attention-based multiple instance learning (ABMIL) network, a classifier each with 128 nodes, a dropout rate of 0.25, and binary cross-entropy loss without class weighting. Models were trained with the AdamW optimizer (initial learning rate 4×10⁻⁵, weight decay 5×10⁻⁵), with the learning rate reduced by a factor (γ) of 0.35 at epochs 5, 12, and 20. The final model, HERA, was retrained on the full cross-validation dataset using this configuration for five epochs, after which training was early-stopped on the internal validation set to mitigate overfitting.

### Development of HERMES

HERMES predicts the 3 molecular VSCC classes: HPV-a, HPV-i/p53wt, and HPV-i/p53abn. For this, we evaluated three architectures that all used H-optimus-1 embeddings as input, which was based on its superior performance and to accommodate experiments on fusion methods with HERA.^16^ A direct multiclass architecture performed comparably to a fusion strategy incorporating HERA’s latent (*m*) embedding and to a multihead HPV/p53 architecture, and was selected for its lower complexity (Table S3). Training hyperparameters were optimized via five-fold cross validation to maximize the macro-AUC, which is the average AUC across all three classes. The optimal configuration used an ABMIL network with an attention module with 256 nodes and a classifier with 128 nodes, a dropout rate of 0.25, and cross-entropy loss with class weighting based on the class distribution (HPV-a: 1.3731, HPV-i/p53wt: 1.2826, HPV-i/p53abn: 0.3443). Models were trained with the AdamW optimizer (initial learning rate 3×10⁻⁵, weight decay 1×10⁻⁵), with the learning rate reduced by a factor (γ) of 0.1 at epochs 5, 15, and 25. The final model, HERMES, was retrained on the full cross-validation dataset using this configuration for eight epochs, after which training was early-stopped on the internal validation set to mitigate overfitting.

### Evaluation metrics

Discriminative performance for both HERA and HERMES was quantified using the area under the receiver operating characteristic curve (AUC), where we used the macro-AUC (average AUC across the classes) for HERMES to give overall discriminative performance. Binary classification performance for HERA was assessed by dichotomizing continuous model probabilities (HERA-scores) into HERA-associated (HERA-a) and HERA-independent (HERA-i) categories using an optimal classification threshold (CT) determined on the internal validation set via Youden’s index.^19^ For the 3-class HERMES model, patients were assigned to one of the 3 molecular subtype (HPV-a, HPV-i/p53wt, or HPV-i/p53abn) based on the maximum predicted class probability (argmax selection rule). To evaluate classification accuracy, we reported sensitivity, specificity, positive predictive value (PPV), negative predictive value (NPV), accompanied by 95% confidence intervals (CIs).

### Blinded pathologist classification

One experienced, expert gynecopathologist (TB) performed the quality assessment and review of all WSIs from the independent testing set (LUMC-I, n=147; UMCG-II, n=139; total n=286). To assess whether model errors reflected morphologically ambiguous cases rather than model-specific failure patterns, this pathologist provided a molecular class prediction based on morphological criteria (eg. basaloid appearance for HPV-a, basal nuclear atypia for HPV-i/p53abn and verrucous growth with minimal atypia for HPV-i/p53wt), acknowledging that these criteria have shown to be imperfect.^20^ For this review the pathologist was blinded to p16, p53 and HPV-PCR results, clinical outcome data, and HERA or HERMES predictions.

### Scoring of growth patterns

The expert pathologist (TB) assigned the growth pattern of the invasive growth into three categories, 1) broad-front infiltration; defined by an endophytic pushing invasive front, 2) spindled-front infiltration; defined by irregular cords or single atypical cells in a so-called spray pattern, often with surrounding desmoplastic stroma and 3) nested invasion; consisted of either large geographic nests or small nests with frequent central keratin pearls.

### Statistical analysis

Statistical tests were two-sided and were considered significant when p < 0.05. Descriptive statistics were applied to assess subgroup proportions and compared using Chi-Square and Fisher’s exact test (adjusted using Holm correction). FIGO stage was compared with the Mann-Whitney U test as an ordinal variable (I–IV). Mann-Whitney U test was used for continuous variables. AUC metrics and classification metrics were evaluated using n=1000 times bootstrapping to report mean with 95% confidence intervals. McNemar’s test was used to compare classification accuracy of ground truth molecular subtypes between HERA/HERMES and the pathologist on discordant cases. Inter-rater agreement between HERA and pathologist classifications was quantified using Cohen’s kappa, and between HERMES and pathologist classifications using Cohen’s unweighted kappa. Recurrence rate, defined as the time in months from diagnosis until date of recurrence (local, groin or distant), was analyzed using the Kaplan–Meier method and groups were compared with the log-rank test.

### Software

HERA and HERMES were developed in Python version 3.12.9 using PyTorch version 2.7.0.^21^ Statistical analysis was performed using R version 4.4.1, R Core Team, Vienna, Austria.^22^ The package OpenSlide was used for pre-processing the WSIs.^23^ Key R packages included survival (version 3.8.6) for recurrence analysis and ggplot2 (version 4.0.3) for data visualization.^24,25^ Figure post-processing was done using Adobe Illustrator (version 30.8; Adobe Inc., San Jose, CA, USA).

## Results

### Cohort clinical and molecular characteristics

Clinical characteristics and molecular feature prevalence are summarized in Table 1 (HPV genotype distributions in Table S4; p53 IHC patterns in Table S5). While the prevalence of HPV-a was similar between the development and independent testing sets (16.5% vs. 16.4%), the distribution of the remaining classes varied (p<0.001), with higher rates of HPV-i/p53wt (17.5% vs. 30.1%) and lower rates of HPV-i/p53abn (66.0% vs. 53.5%) in the testing cohort compared to the model development set.

**Table 1.** Clinical and molecular characteristics of the model development and testing sets. P-values are from Chi-square or Fisher’s exact test as appropriate, excluding missing categories. FIGO stage was compared with the Mann-Whitney U test as an ordinal variable (I–IV), excluding missing values. FIGO, International Federation of Gynecology and Obstetrics; HPV-a, HPV-associated; HPV-i/p53abn, HPV-independent/p53-abnormal; HPV-i/p53wt, HPV-independent/p53-wildtype; IHC, immunohistochemistry; PCR, polymerase chain reaction.

|  | Training | Independent Testing |  |  |  |
| --- | --- | --- | --- | --- | --- |
|  | Model Development (n=644) | LUMC-I (n=147) | UMCG-II (n=139) | Total (n=286) | P-value * |
| **FIGO Stage (n (%))** |  |  |  |  | 0.045 |
| I-II | 149 (23.1%) | 91 (61.9%) | 70 (50.4%) | 161 (56.3%) |  |
| III-IV | 75 (11.7%) | 55 (37.4%) | 69 (49.6%) | 124 (43.4%) |  |
| Missing | 420 (65.2%) | 1 (0.7%) | 0 (0.0%) | 1 (0.3%) |  |
| **Growth Pattern (n (%))** |  |  |  |  | – |
| Broad-front | 0 (0.0%) | 42 (28.6%) | 37 (26.6%) | 79 (27.6%) |  |
| Spindled | 0 (0.0%) | 38 (25.9%) | 35 (25.2%) | 73 (25.5%) |  |
| Nested | 0 (0.0%) | 67 (45.6%) | 67 (48.2%) | 134 (46.9%) |  |
| Not assessed | 644 (100%) | 0 (0.0%) | 0 (0.0%) | 0 (0.0%) |  |
| **Molecular Class** |  |  |  |  | <0.001 |
| HPV-a | 106 (16.5%) | 26 (17.7%) | 21 (15.1%) | 47 (16.4%) |  |
| HPV-i/p53wt | 113 (17.5%) | 43 (29.3%) | 43 (30.9%) | 86 (30.1%) |  |
| HPV-i/p53abn | 425 (66.0%) | 78 (53.1%) | 75 (54.0%) | 153 (53.5%) |  |
| **p16 IHC (n (%))** |  |  |  |  | 0.85 |
| Negative | 534 (82.9%) | 123 (83.7%) | 112 (80.6%) | 235 (82.2%) |  |
| Positive | 110 (17.1%) | 24 (16.3%) | 27 (19.4%) | 51 (17.8%) |  |
| **PCR-HPV Risk (n (%))** |  |  |  |  | 0.49 |
| Negative/Low Risk | 214 (33.2%) | 121 (82.3%) | 118 (84.9%) | 239 (83.6%) |  |
| High Risk | 47 (7.3%) | 25 (17.0%) | 21 (15.1%) | 46 (16.1%) |  |
| Missing | 383 (59.5%) | 1 (0.7%) | 0 (0.0%) | 1 (0.3%) |  |
| **p53 IHC (n (%))** |  |  |  |  | <0.001 |
| Wildtype p53 IHC pattern | 201 (31.2%) | 65 (44.2%) | 56 (40.3%) | 121 (42.3%) |  |
| Mutant-type p53 IHC pattern | 442 (68.6%) | 81 (55.1%) | 80 (57.6%) | 161 (56.3%) |  |
| Missing | 1 (0.2%) | 1 (0.7%) | 3 (2.2%) | 4 (1.4%) |  |

\*Testing for difference between model development and total independent testing set, excluding missing categories.

### Model development and discriminative performance

HERA achieved an AUC of 0.91 (95% CI: 0.86–0.95) in five-fold cross-validation, 0.90 (0.80–0.99) on the internal validation set, and 0.84 (0.75–0.91) on independent testing (model architecture shown in Figure 1; cohort-specific performances in Table S6). HERMES reached macro-average AUCs of 0.77 (0.74–0.80), 0.73 (0.67–0.79), and 0.70 (0.66–0.75) across the same three evaluations. Class-wise AUCs in independent testing were highest for HPV-a (0.80, 0.70–0.88), intermediate for HPV-i/p53abn (0.68, 0.62–0.74), and lowest for HPV-i/p53wt (0.63, 0.56–0.70; Table S6).

**Figure 1.**
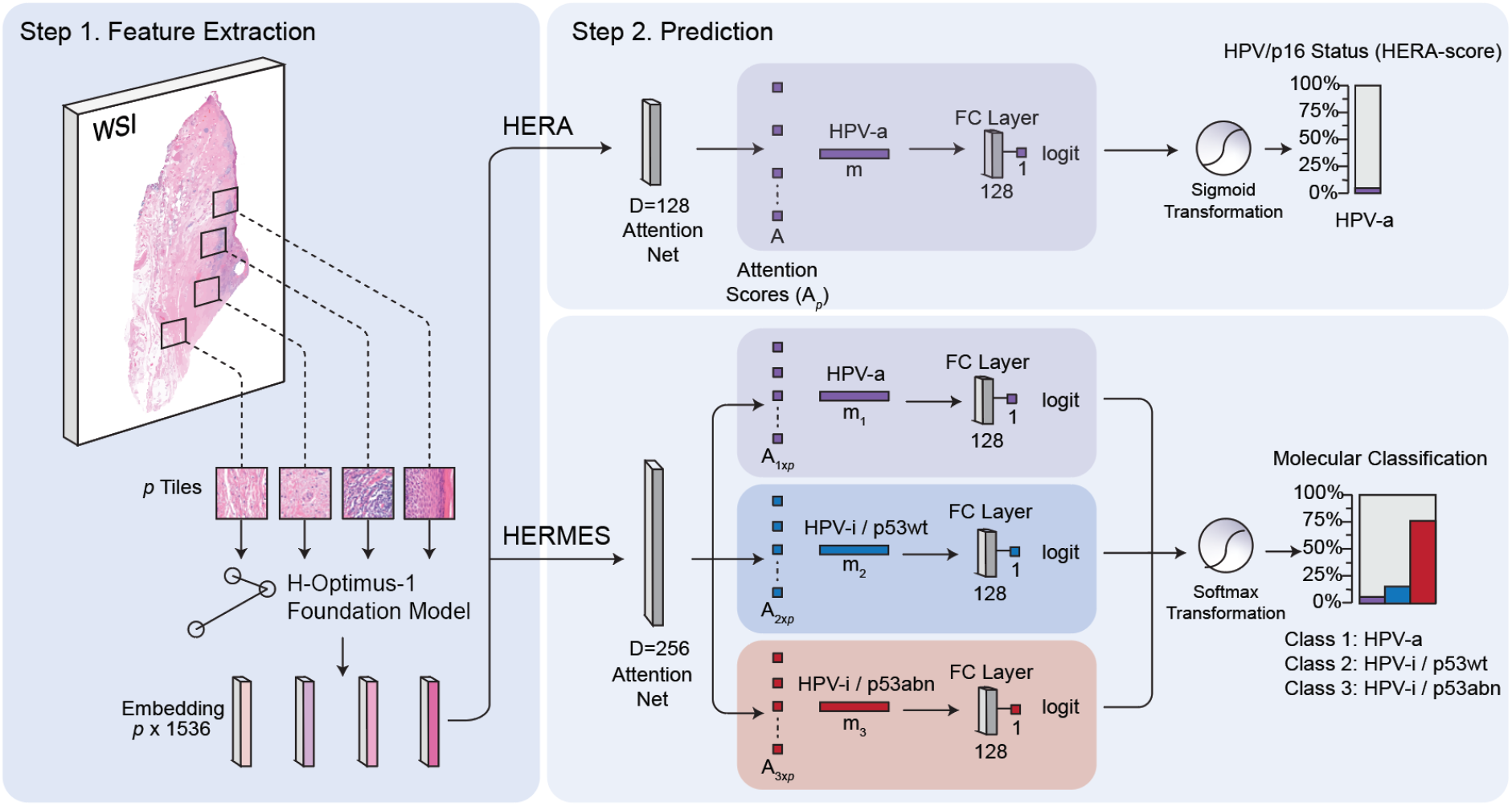
Model architecture of HERA and HERMES for feature extraction and molecular subtype prediction from H&E whole-slide images (WSIs). *Step 1:* Tile-level embeddings are extracted by the H-optimus-1 foundation model from non-overlapping 180-µm tiles at 40x magnification (0.25 µm/pixel) resized to 224 × 224 pixels (≈0.80 µm/px). *Step 2, upper panel:* HERA predicts HPV/p16 status (binary) using an attention-based multiple instance learning (ABMIL) network with a 128-node attention module. Attention scores (A) weight tile embeddings into an aggregated representation (m), which is passed through a 128-node fully connected (FC) layer and a sigmoid classifier to produce a continuous HPV/p16 probability (the HERA-score). *Step 2, lower panel:* HERMES predicts the three molecular subtypes (HPV-associated [HPV-a], HPV-independent/p53-wildtype [HPV-i/p53wt], HPV-independent/p53-abnormal [HPV-i/p53abn]) using three class-specific ABMIL attention modules (256-node attention, 128-node FC) with a softmax classifier producing class probabilities that sum to 1. FC, fully connected layer; MIL, multiple instance learning; WSI, whole-slide image.

### HERA: Classification performance, clinical outcomes, and discordant cases

Classification performance was evaluated using a threshold derived on the internal validation set (HERA-score = 0.1411, Youden index) and applied unchanged to independent testing (Figure 2a–b). This threshold yielded a sensitivity of 0.86 (0.69–1.00) and specificity of 0.93 (0.87–0.97) in internal validation (Figure 2c). In independent testing, specificity remained high (0.93, 0.89–0.96) while sensitivity degraded to 0.68 (0.54–0.80) (Figure 2d; cohort-specific results in Table S7). Negative predictive value was high in both internal validation (0.97 (0.93–1.00)) and independent testing (0.94 (0.90–0.97)).

**Figure 2.**
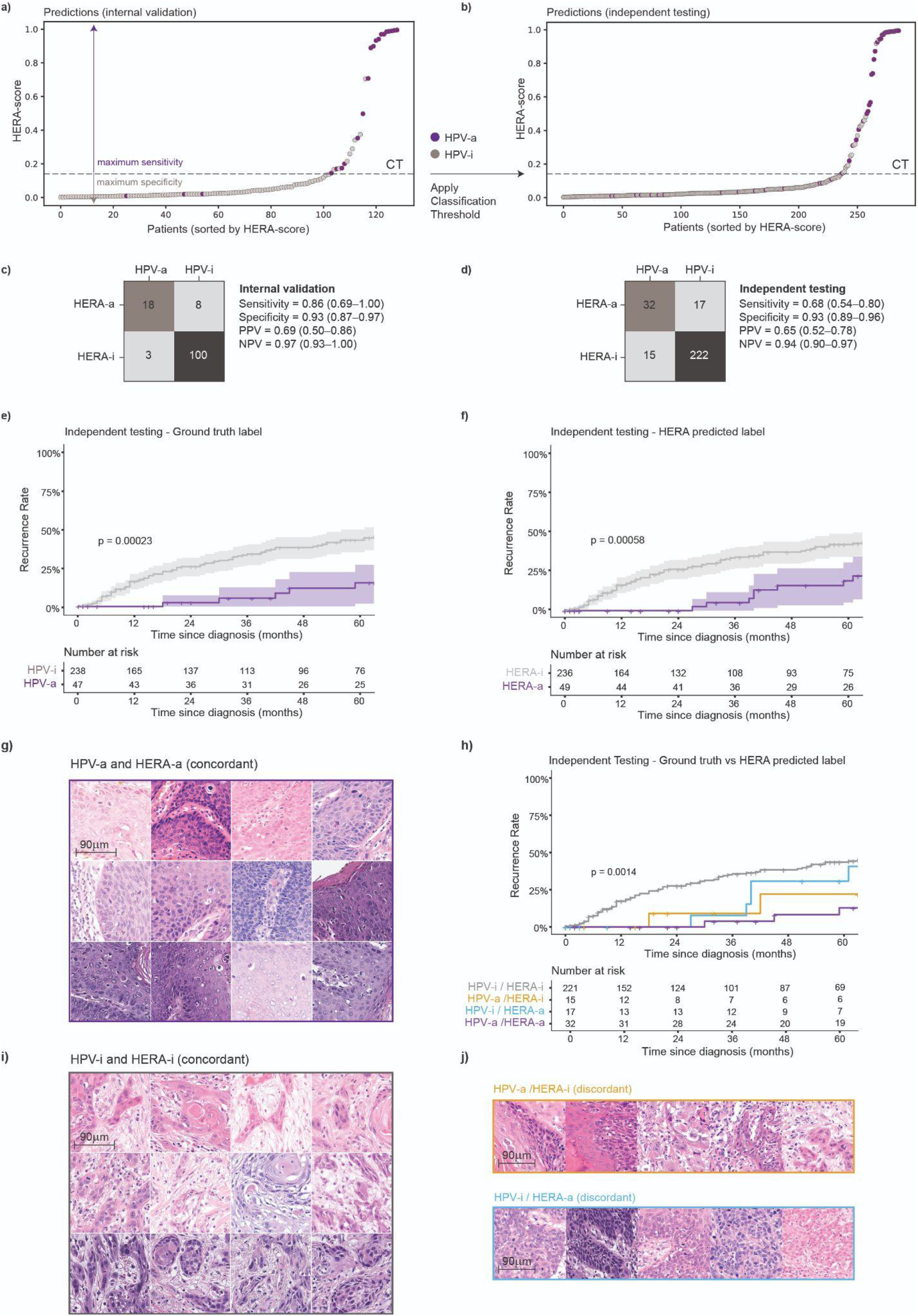
HERA classification performance, recurrence stratification, and morphological characterization. *(a–b)* Distribution of HERA-scores for all patients ranked by score in the internal validation set *(a)* and the independent testing set *(b)*. HPV-a cases are shown in purple, HPV-i cases in grey. The classification threshold (CT; HERA-score = 0.1411), derived on the internal validation set by maximizing Youden’s index, is indicated by a dashed line. *(c–d)* Confusion matrices of HERA predictions against ground-truth labels in the internal validation set *(c)* and the independent testing set *(d)*, with sensitivity, specificity, positive predictive value (PPV), and negative predictive value (NPV) shown alongside. *(e–f)* Kaplan–Meier curves of recurrence rate stratified by ground-truth HPV/p16 label *(e)* and HERA-predicted label *(f)* in the independent testing set (n=285, one case without follow-up excluded). Numbers at risk are shown below each curve. P-values from the log-rank test. *(g)* Representative highly-attended tiles (180×180-µm) from concordantly predicted HPV-a/HERA-a cases. *(h)* Kaplan–Meier curves of recurrence rate for the concordant and discordant prediction groups (HPV-i/HERA-i, HPV-a/HERA-i, HPV-i/HERA-a, HPV-a/HERA-a). Numbers at risk are shown below. P-value from the log-rank test across all four groups. *(i)* Representative highly-attended tiles (180×180-µm) from concordantly predicted HPV-i/HERA-i cases. *(j)* Representative highly-attended tiles (180×180-µm) from discordant cases: HPV-a/HERA-i (yellow border, upper row) and HPV-i/HERA-a (blue border, lower row). CT, classification threshold; HPV-a, HPV-associated; HPV-i, HPV-independent; NPV, negative predictive value; PPV, positive predictive value.

Kaplan–Meier analysis confirmed prognostic separation based on HPV/p16 ground truth status and HERA-predicted groups (Figure 2e–f): 5-year recurrence rate was 15.8% (1.9%–26.9%) for HPV-a versus 42.8% (35.0%–49.6%) for HPV-i VSCC by ground truth (log-rank p<0.001), and 19.6% (5.4%–31.7%) versus 42.1% (34.3%–48.9%) for HERA-a versus HERA-i groups (log-rank p<0.001). Hence, cases predicted as HPV-a by HERA demonstrated significantly fewer recurrences compared to those predicted as HPV-i, indicating that the model captures prognostically relevant morphological features.

To characterize the morphology underlying correct classifications, the ten most highly attended tiles using attention mapping from the twelve most confidently and correctly classified cases per class were reviewed by two expert pathologists (Figure 2g,i). Within the highly attended tiles from HPV-a cases, pathologists identified basaloid cells, koilocytotic-like atypia, and intraepithelial apoptotic bodies. Highly attended tiles from HPV-i tumors displayed invasive tumor nests with keratin pearls surrounded by stromal desmoplasia. Examples of attention maps of correctly classified HPV-a and HPV-i cases are shown in Figure S2.

We examined whether HERA-scores and misclassifications correlated with HPV genotype, discordance in the ground truth label between p16 IHC and HPV-PCR, and growth patterns. There were 32 (11%) misclassifications in total (Table S8). HERA misclassified 15 HPV-a cases as HPV-i (HERA-scores per HPV-type shown in Table S9). The HPV-type 18 tumors received lower HERA-scores (n=4; median 0.029 [0.016–0.052]) than other HPV-type tumors (n=42; median 0.650 [0.118–0.983]; p=0.014) (Figure S3a-b), resulting in four misclassifications. p16−/HPV-a cases (n=7) scored lower HERA-scores than p16+/HPV-a cases (n=40) (0.030 [IQR 0.017–0.072] vs. 0.736 [0.201–0.984], p=0.010; Figure S3c), resulting in six misclassifications (Table S8). Similarly, p16+/HPV-i cases (n=11) scored lower HERA-scores than p16+/HPV-a cases (n=40) (p16+/HPV-i: 0.067 [0.022–0.114] vs. 0.736 [0.201–0.984], p<0.001; Figure S3c), resulting in three misclassifications (Table S8). HERA-scores differed across growth patterns (p<0.001); they were highest in the broad-front group (n=79; HERA-score 0.060 [0.034–0.315]), intermediate in the nested group (n=134; HERA-score 0.031 [0.016–0.066]), and lowest in the spindled group (n=73; HERA-score 0.016 [0.009–0.031]) (Figure S3d). Spindled HPV-a cases were more often misclassified as HPV-i than non-spindled pattern types: 4/4 (100%) versus 4/24 (17%) broad-front and 7/19 (37%) nested (p<0.01).

One expert pathologist classified all cases blinded to ground truth labels into the molecular subtypes, after which we merged the HPV-i/p53wt and HPV-i/p53abn classifications together as HPV-i to compare with HERA classifications (Table S10). The pathologist reached a sensitivity of 57%, a specificity of 86%, PPV of 44%, and a NPV of 91% compared to the ground truth. Inter-rater agreement between the pathologist and HERA was moderate (κ = 0.44 [0.31–0.57]). Of the 15 HPV-a cases misclassified by HERA as HPV-i (32%), the pathologist also misclassified 12 as HPV-i. Among the 17 HPV-i cases misclassified by HERA as HPV-a (7.1%), the pathologist misclassified 6 as HPV-a. When HERA and the pathologist disagreed (50 cases total), HERA predicted the true status more often correctly than the pathologist (n=36 cases (8 HPV-a and 28 HPV-i) compared to 14 (3 HPV-a and 11 HPV-i); p=0.003).

Analyzing the clinical outcomes of discordant cases, we found that HPV-a tumors misclassified as HERA-i trended toward worse outcomes than concordant HPV-a cases (5-year recurrence rate 22.1% (0.0%–45.4%), n=15, vs. 12.8% (0.0%–25.4%), n=32; log-rank p=0.2), while HPV-i tumors misclassified as HERA-a trended toward better outcomes than concordant HPV-i cases (30.8% (0.5%–51.8%), n=17, vs. 43.5% (35.4%–50.6%), n=221; log-rank p=0.2); neither reached statistical significance (Figure 2h; highly-attended tiles shown in Figure 2j).

### HERMES: Classification performance, clinical outcome, and discordant cases

Predicted class probabilities showed a consistent pattern: HPV-a cases received high probabilities for both the HPV-a and HPV-i/p53abn classes, HPV-i/p53wt cases for the HPV-i/p53wt and HPV-i/p53abn classes, and HPV-i/p53abn cases predominantly for the HPV-i/p53abn class (Figure 3a). This was reflected in classification performance: HPV-i/p53abn was called frequently, giving high sensitivity (0.91, 0.86–0.95) but modest PPV (0.63, 0.56–0.69); HPV-i/p53wt was rarely called, giving low sensitivity (0.18, 0.10–0.26); and HPV-a sensitivity was moderate (0.59, 0.44–0.73) with high NPV (0.92, 0.89–0.95) (confusion matrices for independent testing in Figure 3b and for internal validation in Figure S4; cohort-specific classification results in Table S11).

**Figure 3.**
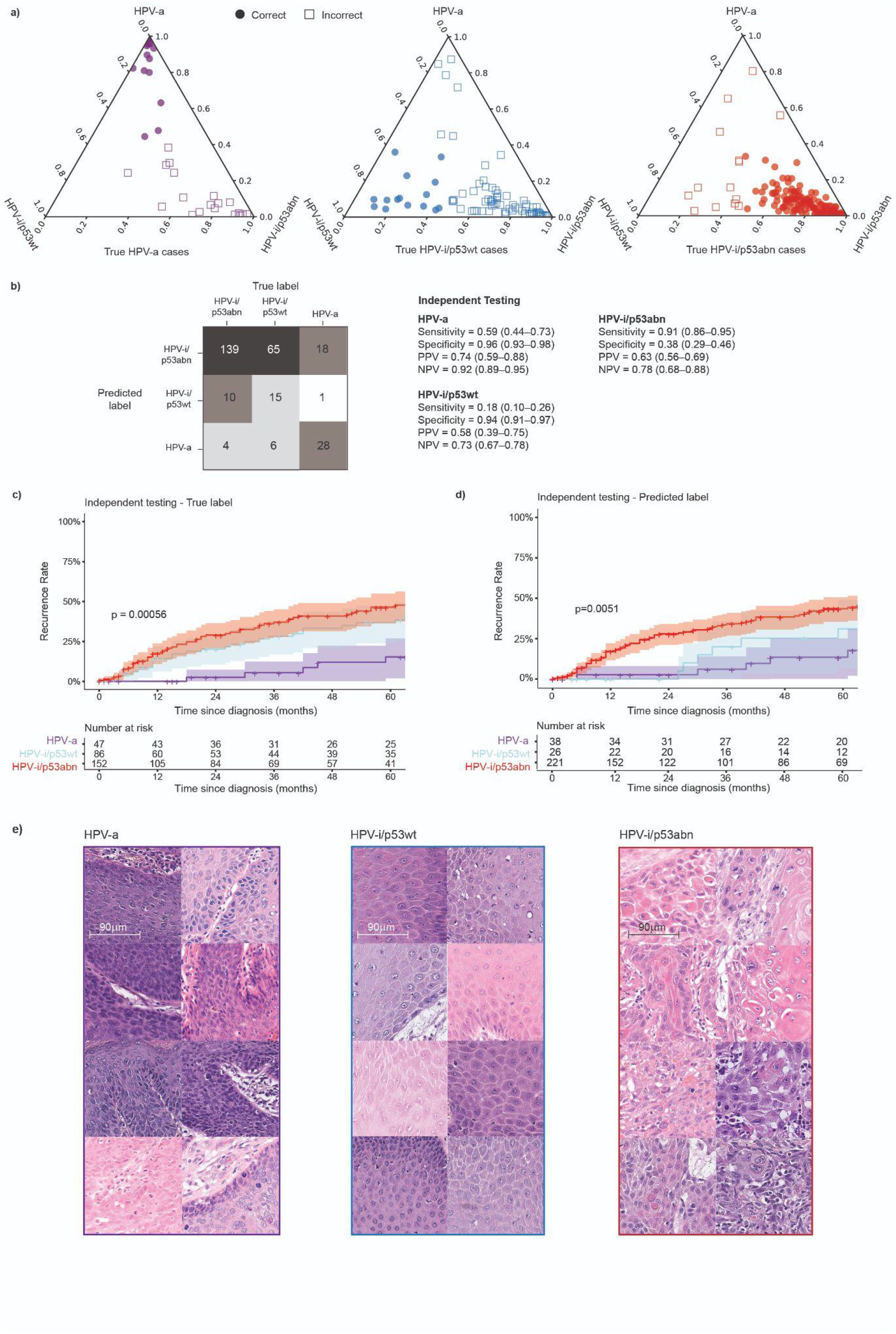
HERMES classification performance, recurrence stratification, and morphological characterization in the independent testing set (n=286). *(a)* Ternary plots showing the distribution of predicted class probabilities for the three true molecular subtype groups: HPV-a cases (n=47, left), HPV-i/p53wt cases (n=86, center), HPV-i/p53abn cases (n=153, right). Correctly classified cases are shown as filled circles; incorrectly classified cases as open squares. Probabilities for HPV-a, HPV-i/p53wt, and HPV-i/p53abn are plotted on the three axes. *(b)* Confusion matrix of HERMES predictions in the independent testing set with class-specific sensitivity, specificity, PPV, and NPV shown alongside. Confusion matrix of internal validation results shown in Figure S4. *(c)* Kaplan–Meier curves of recurrence rate stratified by ground-truth molecular subtype (n=285, one case without follow-up excluded). *(d)* Kaplan–Meier curves of recurrence rate stratified by HERMES-predicted molecular subtype (n=285, one case without follow-up excluded). Numbers at risk are shown below each panel. P-values from log-rank test. *(e)* Representative highly-attended tiles (180×180-µm) from concordantly predicted cases of each molecular subtype: HPV-a (left), HPV-i/p53wt (center), and HPV-i/p53abn (right). HPV-a, HPV-associated; HPV-i/p53abn, HPV-independent/p53-abnormal; HPV-i/p53wt, HPV-independent/p53-wildtype; NPV, negative predictive value; PPV, positive predictive value.

Ground-truth molecular classes showed the expected prognostic hierarchy (5-year recurrence rate: 15.8% (1.9%–27.7%) HPV-a, 35.9% (23.3%–46.5%) HPV-i/p53wt, 46.3% (36.3%–54.8%) HPV-i/p53abn; log-rank p<0.001), and DL-predicted classes reproduced this stratification (14.0% (0.1%–25.9%), 31.1% (6.8%–49.0%), 43.3% (35.2%–50.3%); log-rank p=0.0051) (Figure 3c–d).

Highly attended tiles in the HPV-a class showed the same features identified for HPV-a cases by HERA, which were basaloid cells, koilocytotic-like atypia, and intraepithelial apoptotic bodies. The features of HPV-i tumors identified by HERA, invasive tumor nests with keratin pearls surrounded by stromal desmoplasia, were observed in the HPV-i/p53abn class (Figure 3e). Highly attended tiles in the HPV-i/p53wt class showed well-differentiated keratinocytes with prominent nucleoli, spongiosis, and intercellular bridges. An example attention map of a correctly classified HPV-i/p53wt case is shown in Figure S5.

We examined whether HERMES misclassifications correlated with HPV genotype and growth patterns. HERMES misclassified 19 of 47 (40.4%) HPV-a cases (Figure S6). From these 19 misclassifications, one was predicted as HPV-i/p53wt and was HPV-type 16; the remaining 18 were predicted as HPV-i/p53abn, from which 13 were HPV-type 16, four were HPV-type 18, and one was HPV-type 53. In the HPV-i/p53wt group (n=86), there were 31 cases with broad-front morphology, 37 with nested morphology, and 18 with spindled morphology (Figure S7). 32 out of 37 nested HPV-i/p53wt cases were misclassified, and spindled HPV-i/p53wt cases were more often misclassified than broad-front HPV-i/p53wt cases (18/18 vs. 21/31 misclassifications; adjusted p=0.024).

One expert pathologist classified all cases blinded from ground truth labels into the molecular subtypes. The pathologist did not yield a significantly higher sensitivity than HERMES for the HPV-i/p53wt subtype (27% vs. 18% [10–26%]; p=0.099; Figure S8, Table S12). The pathologist overcalled the HPV-i/p53abn subtype less often than HERMES (specificity 54% vs 38%; p<0.001). We found that the inter-rater agreement between the pathologist and HERMES was fair (unweighted κ = 0.40 [0.30–0.49]). HERMES misclassified 19 HPV-a, 71 HPV-i/p53wt, and 14 HPV-i/p53abn cases; the pathologist also misclassified 14 (74%), 58 (82%), and 8 (57%) of these respective groups (Table S12).

Analyzing the clinical outcomes of cases classified by HERMES stratified by ground truth, we found that among true HPV-i/p53wt cases, the 5-year recurrence rate was 9.1% (0.0%–24.6%) for cases correctly classified by HERMES (n=15) versus 45.2% (29.2%–57.5%) for those misclassified as HPV-i/p53abn (n=65; log-rank p=0.069) (Figure 4a). Among the HPV-i cases predicted as HPV-i/p53abn by HERMES (n=203), the 5-year recurrence rate was 46.0% (35.5%–54.9%) for true HPV-i/p53abn cases (n=138) and 45.2% (29.2%–57.5%) for true HPV-i/p53wt cases (n=65; log-rank p=0.81) (Figure 4b).

**Figure 4.**
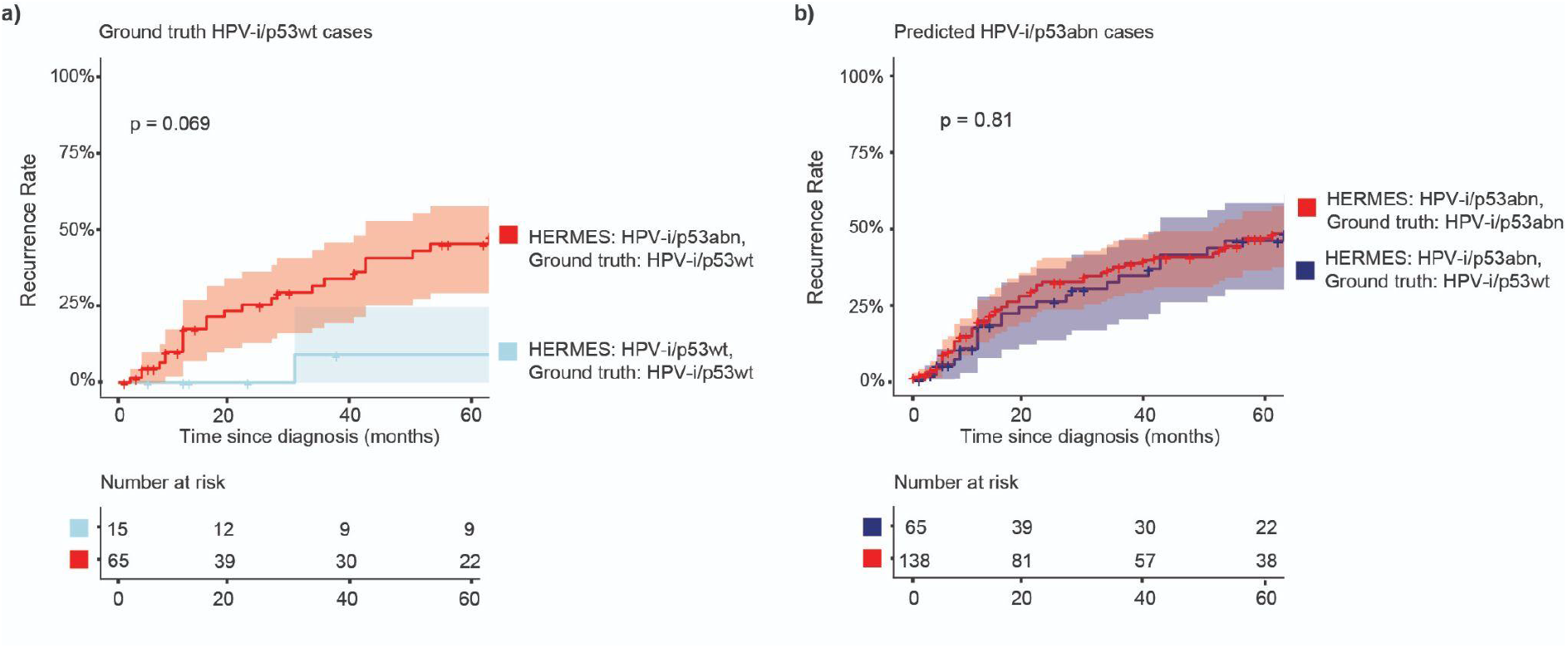
Clinical outcome analysis of concordant and discordant HERMES predictions in HPV-i cases. *(a)* Kaplan–Meier curves of recurrence rate for true HPV-i/p53wt cases (n=80) stratified by HERMES prediction: correctly classified as HPV-i/p53wt (n=15, blue) versus misclassified as HPV-i/p53abn (n=65, red). P-value from log-rank test. *(b)* Kaplan–Meier curves of recurrence rate for HERMES-predicted HPV-i/p53abn cases (n=203) stratified by ground-truth label: true HPV-i/p53abn (n=138, red) versus true HPV-i/p53wt (n=65, blue). P-value from log-rank test. Numbers at risk are shown below each curve. HPV-a, HPV-associated; HPV-i/p53abn, HPV-independent/p53-abnormal; HPV-i/p53wt, HPV-independent/p53-wildtype.

## Discussion

To our knowledge, we here present the first DL models that predict HPV/p16 status and molecular subtypes from H&E-stained whole-slide images of VSCC. The binary model, HERA, distinguished HPV-a from HPV-i VSCC with good discrimination in the independent test set (AUC 0.84, 95%CI 0.75–0.91), despite UMCG-II using an unseen scanner. Stratification by HERA reproduced the prognostic stratification of HPV testing. The multiclass model, HERMES, achieved a macro-AUC of 0.70 (0.66–0.75) in independent testing. HERMES-based stratification preserved the prognostic ordering of the three molecular subtypes.

HERA’s discriminative performance is similar to performances reported in studies for HPV detection in HNSCC, where external validation AUCs range from 0.77 to 0.84, suggesting that HPV-driven squamous carcinogenesis is associated with recognizable morphologic features.^8–10^ Other than HPV prediction, HERA achieved similar performance to MSIntuit predicting microsatellite instability in colorectal cancer (AUC 0.88), but lower performance than EAGLE predicting *EGFR* mutations in lung adenocarcinoma (AUC 0.89) and POLARIX predicting *POLE* mutations in endometrial cancer (AUC 0.95).^26–28^ When comparing model development dataset sizes, MSIntuit had similar training set size (859 WSIs from 434 patients), but EAGLE and POLARIX were trained on substantially larger datasets: 5,174 WSIs from 4,867 patients, and 2,328 WSIs from 2,238 patients, respectively. Although our cohort is large for a rare malignancy (644 WSIs from unique patients), limited training size likely constrains model performance.

Leveraging the clinical outcome data, we found that patients predicted as HPV-a by HERA demonstrated significantly fewer recurrences than those predicted as HPV-i, indicating that the model captures prognostically relevant morphological features. We identified these features using attention mapping, which showed that highly attended tiles from HPV-a tumors displayed basaloid cells, koilocytotic-like atypia, and intraepithelial apoptotic bodies, consistent with the warty and basaloid phenotype of HPV-a VSCC.^20^ Highly attended tiles from HPV-i tumors exhibited prominent keratin pearls surrounded by desmoplastic stroma at the invasive front, in line with the predominantly keratinizing histology of HPV-i VSCC.^20^ Future research could investigate the prognostic value of these morphological features compared to ground truth molecular labels.

In our discordant case analysis, we found evidence that discordances might not originate from HERA’s inability to detect HPV-a cases, but from underlying biological heterogeneity. A distinct subset of HPV-a tumors lacks prototypical HPV-a morphology. We found that false negative calls were enriched for tumors with spindle cell morphology, HPV18 genotype and p16-negative/PCR-positive status. It might be possible that HERA detects transcriptionally active HPV infection rather than latent viral DNA. Moreover, it demonstrates that this subset of cases has an ambiguous morphology within HPV-a VSCC that is challenging to detect by a DL model but motivates further translational studies.

HERMES achieved a macro-AUC of 0.70 (0.66–0.75) in independent testing. Discrimination was good for HPV-a (AUC 0.80) but poor for the two HPV-i subtypes (HPV-i/p53wt AUC 0.63; HPV-i/p53abn AUC 0.68). This reflects the difficulty of separating p53wt from p53abn HPV-i tumors on H&E, resulting in overcalling of HPV-i/p53abn tumors (specificity 0.38) and undercalling of HPV-i/p53wt tumors (sensitivity 0.18). This is likely, at least in part, due to the relative scarcity of HPV-i/p53wt in the training set (n=113, 17.5%). Compared to other multiclass histology-based DL classifiers, the classes predicted least accurately are usually those lacking a distinct morphological phenotype. In endometrial cancer, im4MEC showed the most confusion between the morphologically overlapping *POLE-*mutant and mismatch-repair deficient classes.^29^ In colorectal cancer, imCMS performed worst for consensus molecular subtype (CMS) 3, an epithelial subtype without distinctive stromal or immune features (AUC 0.78 vs 0.83–0.89 for the other subtypes).^30^ As for VSCC, although verruciform growth with minimal atypia has been described in HPV-i/p53wt precursor lesions, morphological criteria for distinguishing p53wt from p53abn in HPV-i tumors are imperfect.^20,31^ Notably, unlike imCMS, where the most prevalent subtype (CMS2) was predicted best, HERMES performed best for HPV-a despite its low prevalence, suggesting that morphological distinctness rather than class size drives performance in VSCC.

Although HERMES had limited discriminative ability for the HPV-i subtypes, its predictions appeared to capture prognostically relevant morphology, dividing true HPV-i/p53wt tumors into two groups that trend towards different outcomes. The cases correctly identified as HPV-i/p53wt had a favorable 5-year recurrence rate (9.1% vs 35.9% for the class as a whole). This group was enriched for broad-front growth and absent among spindled tumors, consistent with the known prognostic value of growth pattern in VSCC.^32,33^ The reviewed highly attended tiles of this group showed well-differentiated keratinocytes with intercellular bridges and spongiosis, often in surface epithelium, similar to the verruciform acanthotic precursor of this subtype.^31,34^ In contrast, the majority classified as HPV-i/p53abn had a 5-year recurrence rate similar to that of true HPV-i/p53abn tumors (45.2% vs 46.3%), a group whose predicted class was characterized by invasive nests with keratin pearls and desmoplastic stroma in concordant cases. Although these differences were not significant in these small subgroups and the morphological observations were qualitative, they suggest that part of HPV-i/p53wt VSCC is phenotypically and clinically similar to HPV-i/p53abn. This may reflect misclassification by p53 IHC, which is an imperfect surrogate for *TP53* mutation status. It is possible that performing *TP53* sequencing could confirm the true biological heterogeneity within this class.^6,7^ Either way, these findings warrant a closer morphological study of HPV-i/p53wt VSCC, as morphology may refine prognosis within this class.

This study has some limitations. First, although the overall cohort is large for a rare malignancy, subgroups remain small, limiting both model training and subgroup analysis. For example, HPV-i/p53wt tumors comprised only 113 cases (17.5%) in model development, which could have limited HERMES’s learning of the HPV-i/p53wt class. The independent test set contained only 12 HPV-a tumors with a non-HPV16 genotype, therefore more extensive validation of differences in HERA-scores between HPV types is required. Second, all cohorts originated from the Netherlands, and the independent test cohorts were scanned on only two scanner platforms. HPV prevalence and genotype distribution vary geographically, and generalizability to populations with different HPV epidemiology, and to transportability to other laboratories, scanners and staining protocols, remains to be established. Third, calibration was beyond the scope of this study and should be evaluated in future work, alongside implementation-tailored classification thresholds for HERA. Fourth, growth pattern assessment and the blinded morphology-based molecular classification were performed by a single expert gynecopathologist. Hence, interobserver variability could not be assessed. Fifth, the morphological characterization of highly attended tiles was a qualitative assessment. Pathologists reviewing tiles with knowledge of the predicted class may preferentially recognize established features, which may introduce confirmation bias. Quantitative feature analyses could be used to confirm the found morpho-molecular correlates. Finally, results of the recurrence analyses of discordant subgroups were based on few cases and should be interpreted as hypothesis-generating.

The two models differ in readiness for research use. HERA is closest: because it requires only a digitized H&E slide, it could assign HPV status to existing VSCC cohorts in which p16 immunohistochemistry and HPV PCR were never performed. These include the GROINSS-V I and II studies, which shaped current guidelines but predate routine HPV assessment.^35,36^ Retrospective molecular testing of such cohorts is often impractical, because tissue blocks may be depleted or unavailable and additional testing is costly. H&E-based annotation would overcome this without consuming tissue and would enable exploratory analyses of HPV-a disease in relation to treatment response and recurrence. Before such use, HERA requires external prospective validation on discriminative and calibration performance across different scanners and in populations beyond the Netherlands. HERMES is not yet ready for this purpose, as it must first be shown to reliably detect the HPV-i/p53wt subtype, which will require cohorts enriched for these tumors. Once validated, both models could thus provide molecular classification for cohorts that currently cannot be studied by subtype.

Beyond cohort annotation, two further directions could bring these models closer to clinical practice. First, it should be investigated whether HPV-a morphology can be predicted from preoperative vulvar biopsies and precursor lesions. In routine practice, p16 and p53 are frequently assessed on the biopsy, and preoperative information increasingly guides surgical planning. Second, because both models already stratify patients by prognosis, they could be extended to predict outcome directly rather than via a molecular subtype as an intermediate step. For both applications, research is required to determine whether predictions are reliable and whether they add clinically meaningful information beyond depth of invasion and other established prognostic factors.

In conclusion, we developed and evaluated the first DL models for VSCC to predict HPV/p16 status and molecular subtypes from routine H&E-stained slides. This work represents an important step toward a pragmatic DL-tool that can predict HPV status for this rare malignancy.

## Data Availability

Generated or analyzed data from this study are all included in the published article. Based on a reasonable request, it is available from the corresponding author.

https://github.com/AIRMEC/HPV-AI

## Acknowledgements

We would like to thank Natalja ter Haar and Tessa Rutten for their technical support. We acknowledge the SHARK team, managing the computational cluster of the LUMC, for their technical support.

## Ethics Approval

This retrospective, multicenter cohort study was reviewed and approved by the medical ethical review board of the University Medical Center Groningen (METc 2024/147), and waived the requirement for additional informed consent. The study was additionally approved by the medical ethics committee of the Leiden University Medical Center (B16.024). All research activities were conducted in accordance with the Dutch code of Conduct for Health Research and the principles of the Declaration of Helsinki.

## Author Contribution Statement

JB, TB, WK conceived the study. NB, WK, MO, JB, TB participated in designing the study. MO, JB, NH, TB supervised the study. NB, WK, KL, KK, LN, AFV, NG, LCD collected the data. NB, WK, AIRMEC members contributed to code writing. NB contributed to model training. NB and WK contributed to data analysis. NB and WK designed the figures and drafted the manuscript. NB, WK, AIRMEC, NH, MO, JB, TB revised the manuscript. NB, AIRMEC prepared the code repository. AIRMEC, NH, MO, JB, TB contributed to expert review and data interpretation. All authors had access to the final version of the manuscript and could read and approve. Due to general data protection regulations, verification and access to raw data was limited to NB, WK, JB, TB. AIRMEC is a collaborative research consortium; its contributing members are listed in “AIRMEC Consortium Members”. All authors agreed on submitting the manuscript.

## AIRMEC Consortium Members

Lydia A. Schoenpflug^1,2^, Jurriaan Barkey Wolf^3^, Maxime W. Lafarge^1,2^, Nanda Horeweg^4^, Viktor H. Koelzer^1,2^

¹Institute of Medical Genetics and Pathology, University Hospital Basel, Basel, Switzerland

²Department of Biomedical Engineering, University of Basel, Basel, Switzerland

³Department of Pathology, Leiden University Medical Center, Leiden, the Netherlands

⁴Department of Radiation Oncology, Leiden University Medical Center, Leiden, the Netherlands

## Code Availability Statement

Code used in this study is available via GitHub at https://github.com/AIRMEC/HPV-AI.

