## Supplementary Information for "Predicting HPV/p16 Status and Molecular Subtypes from Histopathology Slides using Deep Learning in Vulvar Squamous Cell Carcinoma"

### Supplement Information


**Data** **2**

Patient Inclusion 2

Slide selection and scanning 2

**Hyperparameter Optimization and Architecture Selection** **3**

HERA 3

Multiclass Architecture Ablation Studies 3

**HPV types per cohort** **4**

**p53 patterns per cohort** **5**

**Discrimination results** **6**

**HERA: Classification results** **7**

**HERA: Attention mapping** **8**

**HERA: Misclassification analysis** **9**

**HERMES: Classification results** **12**

**HERMES: Attention mapping** **14**

**HERMES: Misclassification Analysis** **15**

### Data

#### Patient Inclusion

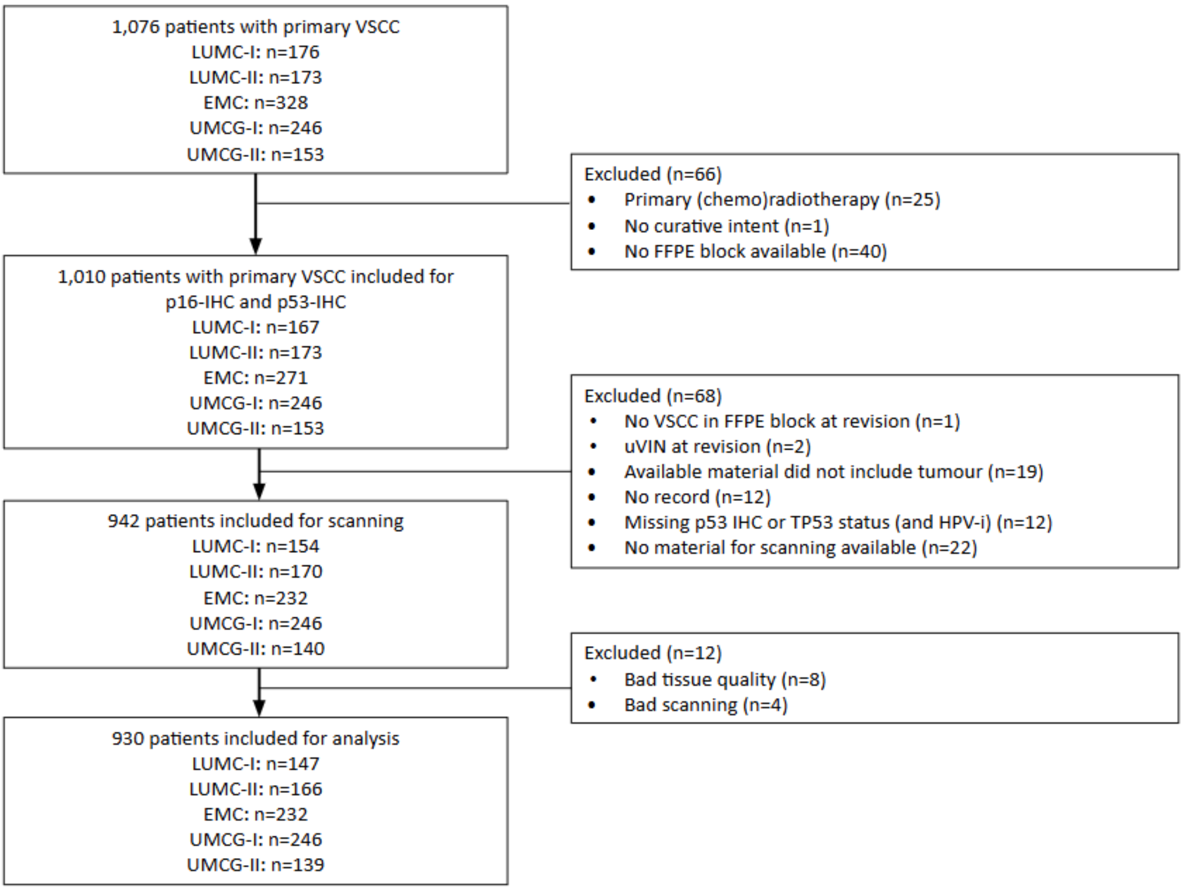


**Figure S1.** CONSORT Inclusion flowchart. FFPE = formalin-fixed paraffin-embedded, IHC = immunohistochemistry, VSCC = vulvar squamous cell carcinoma.

#### Slide selection and scanning

**Table S1.** Scanner information.

| Cohort | Scanner | Objective magnification | mpxx/mpxy [µm per pixel] |
|---|---|---|---|
| EMC | 3D-Histech P480 | 40x | 0.194/0.194 |
| LUMC-I | 3D-Histech P480 | 40x | 0.194/0.194 |
| LUMC-II | 3D-Histech P480 | 40x | 0.194/0.194 |
| UMCG-I | Philips IntelliSite Ultra Fast Scanner (UFS) | 40x | 0.25/0.25 |
| UMCG-II | Aperio ScanScope GL | 40x | 0.2302/0.2302 |

### Hyperparameter Optimization and Architecture Selection

#### HERA

**Table S2.** Cross validation results of foundation models averaged over hyperparameter search for HERA.

| Foundation model | N. of hyperparameter searches | Mean | STD | Min | Max |
|---|---|---|---|---|---|
| H-optimus-1 | 10 | 0.889 | 0.030 | 0.821 | 0.913 |
| UNI2-h | 13 | 0.886 | 0.025 | 0.823 | 0.905 |
| Virchow2 | 6 | 0.873 | 0.015 | 0.846 | 0.887 |

#### Multiclass Architecture Ablation Studies

**Table S3.** Cross validation results per prediction class of the best model architectures, in units of fold means with standard deviation across folds.

| Model | HPV-a | HPV-i/p53wt | HPV-i/p53abn | macro |
|---|---|---|---|---|
| HERA | 0.913 ± 0.049 | Not Applicable | Not Applicable | Not Applicable |
| HERMES (Multiclass model) | 0.879 ± 0.075 | 0.665 ± 0.043 | 0.768 ± 0.036 | 0.773 ± 0.044 |
| “m” Fusion model | 0.907 ± 0.060 | 0.647 ± 0.070 | 0.767 ± 0.032 | 0.779 ± 0.038 |
| Multihead model | 0.891 ± 0.090 | Not Applicable | 0.650 ± 0.113 | 0.739 ± 0.067 |

The three approaches were: (1) a multiclass architecture using three class-specific attention maps, each producing a separate "m" embedding, combined via a softmax output; (2) an "m" fusion strategy in which a single "m" embedding from the multiclass model's attention module was concatenated with the single "m" embedding from the locked, pretrained HERA's attention module, rather than using three class-specific embeddings (cross-validation splits were identical between the binary and multi subtyping model to prevent data leakage) and (3) a multihead framework using a shared attention network with independent binary classifier heads for HPV status (associated/independent) and p53 abnormality (wildtype/abnormal). The direct multiclass and "m" fusion architectures achieved comparable performance; given its lower complexity, we selected the direct multiclass architecture for all downstream analyses.

### HPV types per cohort

**Table S4.** HPV types of the model development and testing sets in HPV-a cases.

| | **Training** | **Independent Testing** | | |
|---|---|---|---|---|
| | **Model Development (n=106)** | **LUMC-I (n=26)** | **UMCG-II (n=21)** | **Total (n=47)** |
| **HPV Type (n (%))** | | | | |
| Type 16 | 42 (39.6%) | 17 (65.4%) | 17 (81%) | 34 (72.3%) |
| Type 18 | 2 (1.9%) | 3 (11.5%) | 1 (4.8%) | 4 (8.5%) |
| Type 31 | 1 (0.9%) | 0 (0%) | 0 (0%) | 0 (0%) |
| Type 33 | 1 (0.9%) | 5 (19.2%) | 1 (4.8%) | 6 (12.8%) |
| Type 52 | 0 (0%) | 0 (0%) | 1 (4.8%) | 1 (2.1%) |
| Type 53 | 0 (0%) | 0 (0%) | 1 (4.8%) | 1 (2.1%) |
| Type 58 | 1 (0.9%) | 0 (0%) | 0 (0%) | 0 (0%) |
| Missing | 59 (55.7%) | 1 (3.8%) | 0 (0%) | 1 (2.1%) |

### p53 patterns per cohort

**Table S5.** p53 patterns of the model development and testing sets. *These cases were included as HPV-a with unknown p53 pattern status.

| | Training | Independent Testing | | |
|---|---|---|---|---|
| | **Model Development (n=644)** | **LUMC-I (n=147)** | **UMCG-II (n=139)** | **Total (n=286)** |
| **p53 wildtype** | | | | |
| Scattered | 24 (3.7%) | 0 (0.0%) | 0 (0.0%) | 0 (0.0%) |
| Mid-epithelial with basal sparing | 8 (1.2%) | 0 (0.0%) | 0 (0.0%) | 0 (0.0%) |
| Wildtype, pattern not recorded | 169 (26.2%) | 65 (44.2%) | 56 (40.3%) | 121 (42.3%) |
| **p53 abnormal** | | | | |
| Basal overexpression | 46 (7.1%) | 6 (4.1%) | 0 (0.0%) | 6 (2.1%) |
| Parabasal/diffuse overexpression | 301 (46.7%) | 70 (47.6%) | 0 (0.0%) | 70 (24.5%) |
| Null (absent) | 82 (12.7%) | 5 (3.4%) | 2 (1.4%) | 7 (2.4%) |
| Cytoplasmatic | 6 (0.9%) | 0 (0.0%) | 0 (0.0%) | 0 (0.0%) |
| Abnormal, pattern not recorded | 7 (1.1%) | 0 (0.0%) | 3 (2.2%) | 4 (1.4%) |
| **Unknown*** | 1 (0.2%) | 1 (0.7%) | 3 (2.2%) | 4 (1.4%) |
| **Total wildtype/abnormal** | 201/442 | 65/81 | 56/80 | 121/161 |

### Discrimination results

**Table S6**. AUCs in training, internal validation and independent testing (LUMC-I and UMCG-II) for HERA and HERMES. Confidence intervals are computed using bootstrapping.

| | HERA | HERMES | | | |
|---|---|---|---|---|---|
| **Cohort** | **HPV-a** | **HPV-a** | **HPV-i/p53wt** | **HPV-i/p53abn** | **Macro** |
| Cross-Validation | 0.91 (0.86–0.95) | 0.89 (0.83–0.93) | 0.67 (0.61–0.73) | 0.76 (0.72–0.81) | 0.77 (0.74–0.80) |
| Internal Validation Set | 0.90 (0.80–0.99) | 0.88 (0.77–0.97) | 0.59 (0.46–0.72) | 0.72 (0.62–0.81) | 0.73 (0.67–0.79) |
| Independent Testing Set | 0.84 (0.75–0.91) | 0.80 (0.70–0.88) | 0.63 (0.56–0.70) | 0.68 (0.62–0.74) | 0.70 (0.66–0.75) |
| - LUMC-I | 0.82 (0.72–0.91) | 0.76 (0.62–0.88) | 0.61 (0.51–0.71) | 0.65 (0.56–0.74) | 0.67 (0.61–0.73) |
| - UMCG-II | 0.87 (0.73–0.99) | 0.89 (0.77–0.98) | 0.68 (0.58–0.78) | 0.75 (0.66–0.83) | 0.77 (0.71–0.82) |

### HERA: Classification results

**Table S7**. Classification performances of HERA for HPV-a using sensitivity, specificity, positive predictive value (PPV), negative predictive value (NPV).

| HERA | | | | |
|---|---|---|---|---|
| **Cohort** | **Sens (95%CI)** | **Spec (95%CI)** | **PPV (95%CI)** | **NPV (95%CI)** |
| | **HPV-a** | | | |
| Internal Validation Set | 0.86 (0.69–1.0) | 0.93 (0.87–0.97) | 0.69 (0.50–0.86) | 0.97 (0.93–1.0) |
| Independent Testing Set | 0.68 (0.54–0.80) | 0.93 (0.89–0.96) | 0.65 (0.52–0.78) | 0.94 (0.90–0.97) |
| - LUMC-I | 0.58 (0.39–0.77) | 0.90 (0.85–0.95) | 0.56 (0.36–0.75) | 0.91 (0.85–0.96) |
| - UMCG-II | 0.81 (0.62–0.95) | 0.96 (0.92–0.99) | 0.77 (0.59–0.94) | 0.97 (0.93–0.99) |

### HERA: Attention mapping

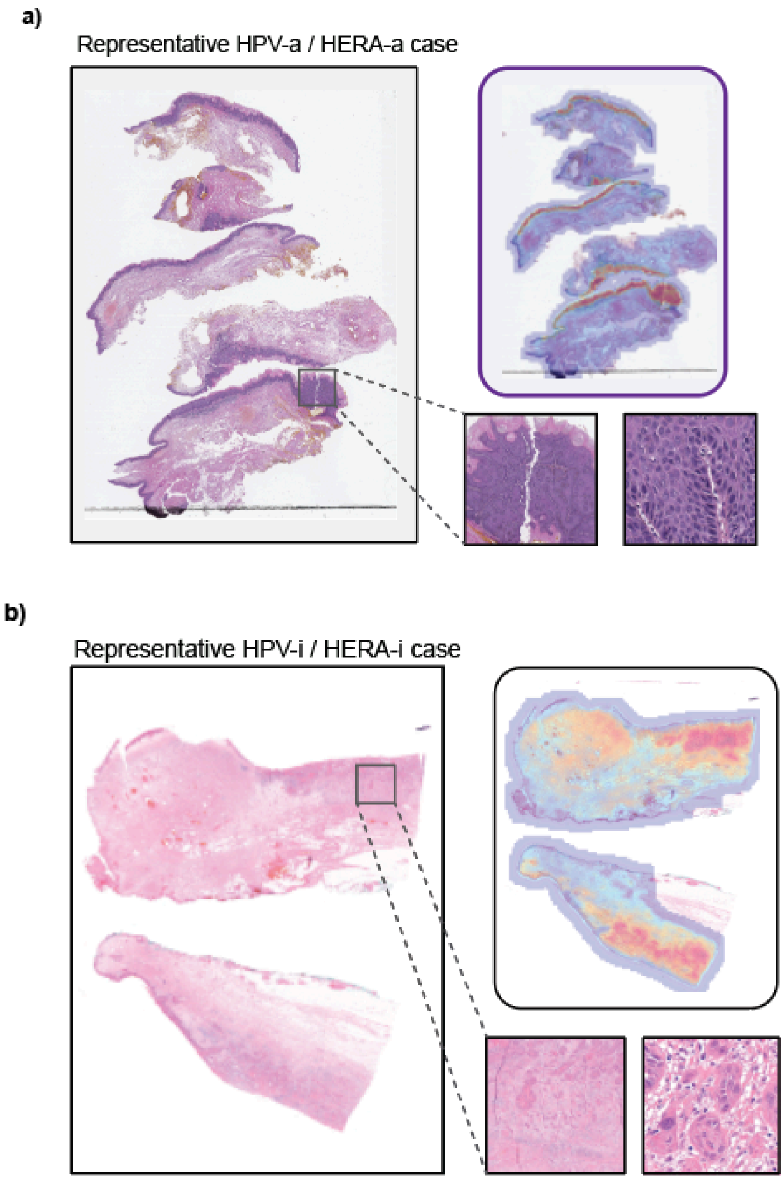


**Figure S2.** Examples of attention maps computed by the ABMIL network for a representative (a) HPV-a/HERA-a case and a (b) HPV-i/HERA-i case.

### HERA: Misclassification analysis

**Table S8.** Misclassification overview (HERA).

| Ground truth | HERA classification | HPV type | p16 IHC | Growth pattern |
|---|---|---|---|---|
| HPV-i | HPV-a | Not detected | Negative | Spindled |
| HPV-i | HPV-a | Not detected | Negative | Broad-front |
| HPV-i | HPV-a | Not detected | Negative | Broad-front |
| HPV-i | HPV-a | Not detected | Negative | Spindled |
| HPV-i | HPV-a | Not detected | Negative | Spindled |
| HPV-i | HPV-a | Not detected | Negative | Nested |
| HPV-i | HPV-a | Not detected | Negative | Broad-front |
| HPV-i | HPV-a | Not detected | Positive | Broad-front |
| HPV-i | HPV-a | Not detected | Negative | Nested |
| HPV-i | HPV-a | Not detected | Negative | Nested |
| HPV-i | HPV-a | Not detected | Negative | Broad-front |
| HPV-i | HPV-a | Not detected | Negative | Nested |
| HPV-a | HPV-i | Type 16 | Negative | Nested |
| HPV-a | HPV-i | Type 16 | Positive | Nested |
| HPV-a | HPV-i | Type 16 | Negative | Broad-front |
| HPV-a | HPV-i | Type 16 | Positive | Nested |
| HPV-a | HPV-i | Type 18 | Positive | Broad-front |
| HPV-a | HPV-i | Type 16 | Negative | Spindled |
| HPV-a | HPV-i | Type 16 | Positive | Nested |
| HPV-a | HPV-i | Type 18 | Positive | Nested |
| HPV-a | HPV-i | Type 16 | Negative | Nested |
| HPV-a | HPV-i | Type 18 | Positive | Broad-front |
| HPV-a | HPV-i | Type 16 | Positive | Nested |
| HPV-i | HPV-a | Not detected | Negative | Nested |
| HPV-a | HPV-i | Type 16 | Negative | Spindled |
| HPV-i | HPV-a | Not detected | Negative | Nested |
| HPV-i | HPV-a | Not detected | Positive | Nested |
| HPV-a | HPV-i | Type 16 | Positive | Spindled |
| HPV-a | HPV-i | Type 18 | Positive | Spindled |

| | | | | |
|---|---|---|---|---|
| HPV-i | HPV-a | Not detected | Positive | Nested |
| HPV-i | HPV-a | Not detected | Negative | Broad-front |
| HPV-a | HPV-i | Type 53 | Negative | Broad-front |

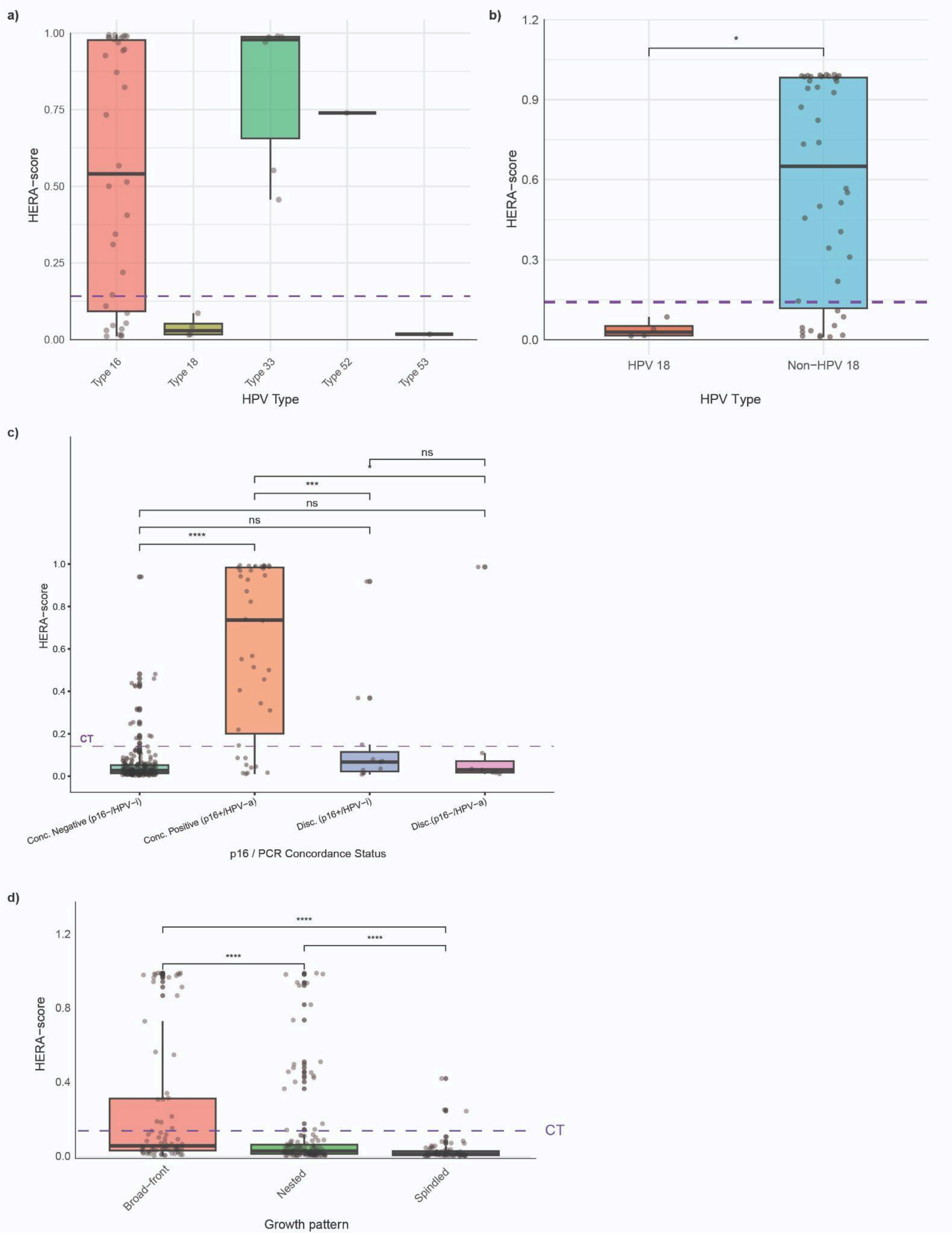


**Figure S3.** Distributions of HERA-scores stratified by tumor characteristics in the independent testing set. *(a)* HERA-scores by HPV genotype among HPV-a cases (n=46 with known genotype). *(b)* HERA-scores by HPV-18 and non-HPV-18 (all HPV-a). The purple dashed line is the classification threshold. *(c)* HERA-scores stratified by p16/PCR concordance status. *(d)* HERA-scores stratified by invasive growth pattern (broad-front, n=79; nested, n=134; spindled, n=73). CT, classification threshold (dashed line).

**Table S9.** HERA-score distributions per HPV-type in HPV-a cases.

| HPV type | N | Median (IQR) |
|---|---|---|
| Type 16 | 34 | 0.540 (0.092 - 0.977) |
| Type 18 | 4 | 0.029 (0.016 - 0.052) |
| Type 33 | 6 | 0.979 (0.656 - 0.988) |
| Type 52 | 1 | 0.739 |
| Type 53 | 1 | 0.017 |

**Table S10**. Counts of HERA prediction groups stratified by ground truth status and pathologist classification.

| Ground truth | HERA | Pathologist | Count |
|---|---|---|---|
| HPV-a | HPV-a | HPV-a | 24 |
| HPV-a | HPV-a | HPV-i | 8 |
| HPV-a | HPV-i | HPV-a | 3 |
| HPV-a | HPV-i | HPV-i | 12 |
| HPV-i | HPV-i | HPV-i | 194 |
| HPV-i | HPV-i | HPV-a | 28 |
| HPV-i | HPV-a | HPV-i | 11 |
| HPV-i | HPV-a | HPV-a | 6 |

### HERMES: Classification results

**Table S11**. Classification performances of HERMES for all classes using sensitivity, specificity, positive predictive value (PPV), negative predictive value (NPV).

| HERMES | | | | |
|---|---|---|---|---|
| **Cohort** | **Sens (95%CI)** | **Spec (95%CI)** | **PPV (95%CI)** | **NPV (95%CI)** |
| | **HPV-a** | | | |
| Internal validation set | 0.72 (0.52–0.90) | 0.97 (0.94–1.0) | 0.83 (0.65–1.0) | 0.95 (0.90–0.98) |
| Independent Testing Set | 0.59 (0.44–0.73) | 0.96 (0.93–0.98) | 0.74 (0.59–0.88) | 0.92 (0.89–0.95) |
| - LUMC-I | 0.54 (0.36–0.74) | 0.95 (0.91–0.98) | 0.70 (0.5–0.89) | 0.91 (0.85–0.95) |
| - UMCG-II | 0.67 (0.47–0.87) | 0.97 (0.93–0.99) | 0.78 (0.56–0.95) | 0.94 (0.90–0.98) |
| | **HPV-i/p53wt** | | | |
| Internal validation set | 0.09 (0.00–0.23) | 0.97 (0.94–1.0) | 0.39 (0.0–1.0) | 0.84 (0.77–0.90) |
| Independent Testing Set | 0.18 (0.10–0.26) | 0.94 (0.91–0.97) | 0.58 (0.39–0.75) | 0.73 (0.67–0.78) |
| - LUMC-I | 0.09 (0.02–0.19) | 0.94 (0.90–0.98) | 0.40 (0.10–0.75) | 0.72 (0.64–0.79) |
| - UMCG-II | 0.25 (0.13–0.38) | 0.95 (0.90–0.99) | 0.68 (0.44–0.92) | 0.74 (0.67–0.81) |
| | **HPV-i/p53abn** | | | |
| Internal validation set | 0.93 (0.87–0.98) | 0.40 (0.24–0.55) | 0.75 (0.67–0.84) | 0.74 (0.55–0.90) |
| Independent Testing Set | 0.91 (0.86–0.95) | 0.38 (0.29–0.46) | 0.63 (0.56–0.69) | 0.78 (0.68–0.88) |
| - LUMC-I | 0.89 (0.81–0.95) | 0.31 (0.20–0.41) | 0.59 (0.50–0.68) | 0.70 (0.53–0.86) |
| - UMCG-II | 0.93 (0.87–0.99) | 0.46 (0.33–0.57) | 0.67 (0.58–0.76) | 0.85 (0.71–0.97) |

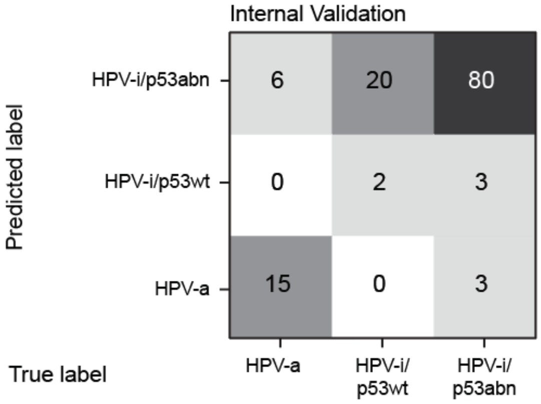


**Figure S4.** Confusion matrix comparing HERMES predicted classes (“predicted label”) and ground truth (“true label”) labels of the internal validation set.

### HERMES: Attention mapping

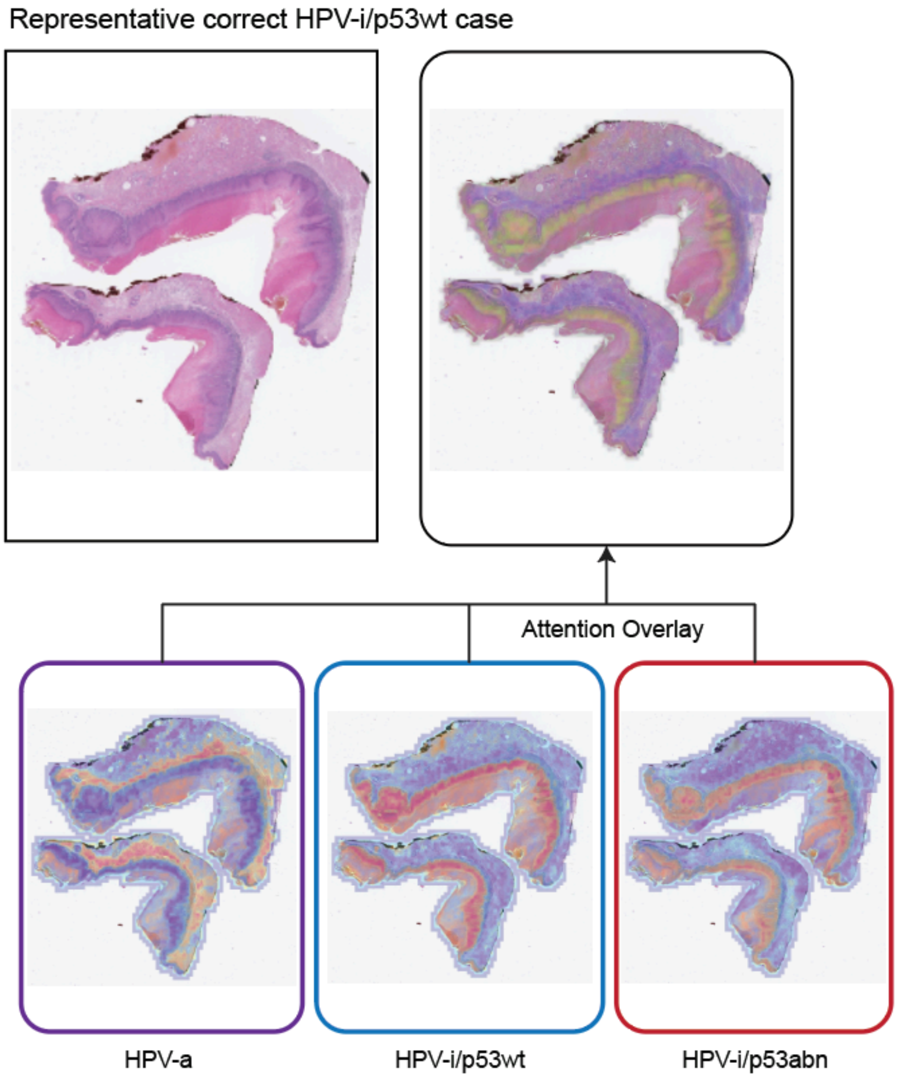


**Figure S5.** Class-specific attention maps for a representative correctly classified HPV-i/p53wt case. Attention overlays are shown separately for each of the three HERMES ABMIL heads (HPV-a, left; HPV-i/p53wt, center; HPV-i/p53abn, right). In this case, the HPV-i/p53wt head localizes predominantly to surface epithelial regions, consistent with the morphological features identified in the image gallery (Figure 3e). ABMIL, attention-based multiple instance learning; HPV-i/p53wt, HPV-independent/p53-wildtype.

### HERMES: Misclassification Analysis

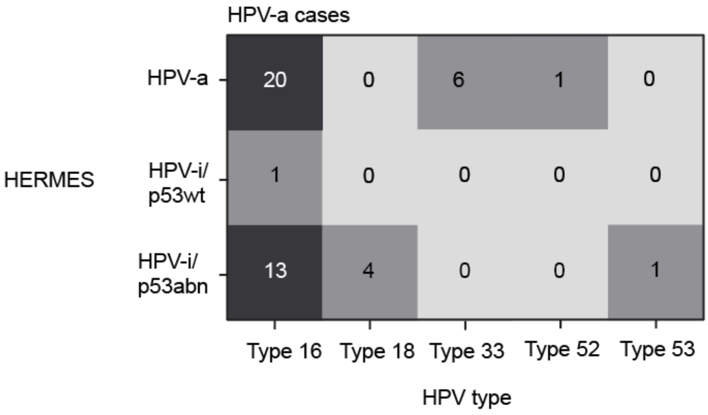


**Figure S6.** Confusion matrices of HERMES classifications compared with HPV-types (ground truth HPV-a cases).

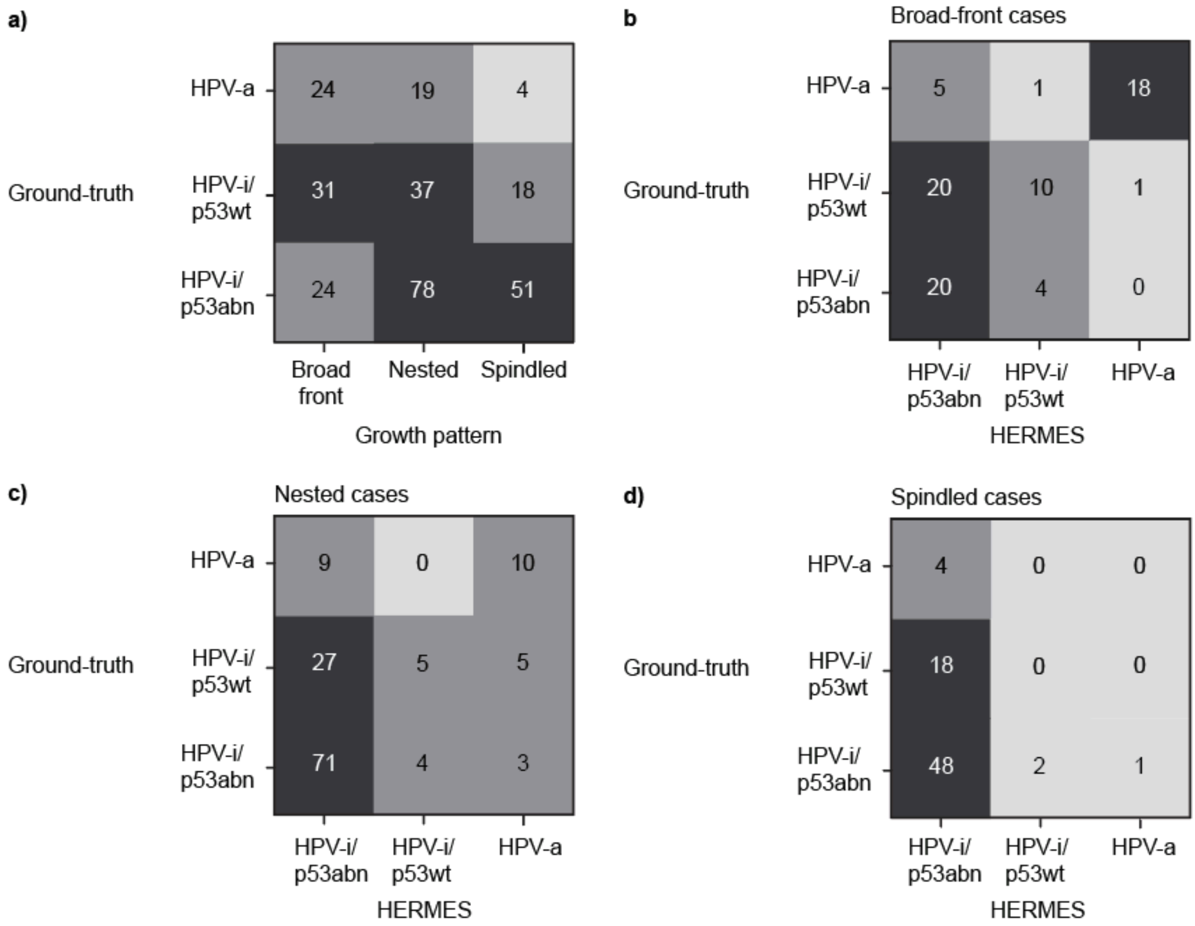


**Figure S7.** Confusion matrices comparing (a) ground truth and growth pattern, (b) ground truth and HERMES classifications in broad-front cases, (c) ground truth and HERMES classifications in nested cases, (d) ground truth and HERMES classifications in spindled cases.

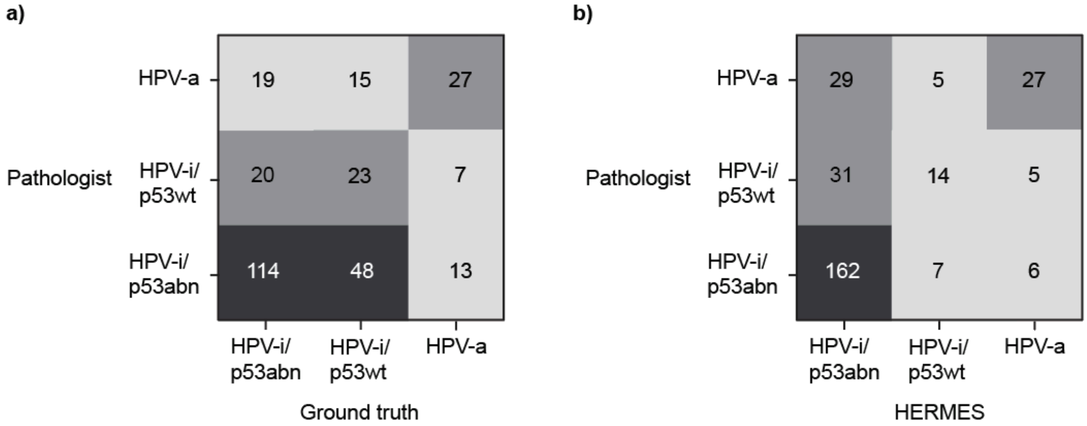


**Figure S8.** Confusion matrices of (a) pathologist classification against ground truth label and (b) pathologist classification against HERMES classification.

**Table S12**. Counts of HERMES prediction groups stratified by ground truth status and pathologist classification.

| Ground truth | HERMES | Pathologist | Count |
|---|---|---|---|
| HPV-a | HPV-a | HPV-i/p53abn | 3 |
| HPV-a | HPV-a | HPV-i/p53wt | 3 |
| HPV-a | HPV-a | HPV-a | 22 |
| HPV-a | HPV-i/p53wt | HPV-a | 1 |
| HPV-a | HPV-i/p53abn | HPV-i/p53abn | 10 |
| HPV-a | HPV-i/p53abn | HPV-i/p53wt | 4 |
| HPV-a | HPV-i/p53abn | HPV-a | 4 |
| HPV-i/p53wt | HPV-a | HPV-i/p53wt | 2 |
| HPV-i/p53wt | HPV-a | HPV-a | 3 |
| HPV-i/p53wt | HPV-a | HPV-i/p53abn | 1 |
| HPV-i/p53wt | HPV-i/p53wt | HPV-i/p53abn | 3 |
| HPV-i/p53wt | HPV-i/p53wt | HPV-i/p53wt | 10 |
| HPV-i/p53wt | HPV-i/p53wt | HPV-a | 2 |
| HPV-i/p53wt | HPV-i/p53abn | HPV-i/p53abn | 44 |
| HPV-i/p53wt | HPV-i/p53abn | HPV-i/p53wt | 11 |
| HPV-i/p53wt | HPV-i/p53abn | HPV-a | 10 |
| HPV-i/p53abn | HPV-a | HPV-i/p53abn | 2 |
| HPV-i/p53abn | HPV-a | HPV-a | 2 |
| HPV-i/p53abn | HPV-i/p53wt | HPV-i/p53abn | 4 |
| HPV-i/p53abn | HPV-i/p53wt | HPV-i/p53wt | 4 |
| HPV-i/p53abn | HPV-i/p53wt | HPV-a | 2 |
| HPV-i/p53abn | HPV-i/p53abn | HPV-i/p53abn | 108 |
| HPV-i/p53abn | HPV-i/p53abn | HPV-i/p53wt | 16 |
| HPV-i/p53abn | HPV-i/p53abn | HPV-a | 15 |
